# Shorter serial intervals in SARS-CoV-2 cases with Omicron BA.1 variant compared to Delta variant in the Netherlands, 13 – 26 December 2021

**DOI:** 10.1101/2022.01.18.22269217

**Authors:** Jantien A. Backer, Dirk Eggink, Stijn P. Andeweg, Irene K. Veldhuijzen, Noortje van Maarseveen, Klaas Vermaas, Boris Vlaemynck, Raf Schepers, Susan van den Hof, Chantal B.E.M. Reusken, Jacco Wallinga

## Abstract

The SARS-CoV-2 Omicron variant has a growth advantage over the Delta variant, due to higher transmissibility, immune evasion, or a shorter serial interval. Using S-gene target failure (SGTF) as indication for Omicron BA.1, we identify 908 SGTF and 1621 non-SGTF serial intervals in the same period. Within households, we find that the mean serial interval for SGTF cases is 0.2-0.6 days shorter than for non-SGTF cases. This suggests that the growth advantage of Omicron is partly due to a shorter serial interval.

---

The SARS-CoV-2 Pango lineage B.1.1.529, also known as the Omicron variant, was first reported by South Africa on 24 November 2021, and designated by the WHO as a variant of concern on 26 November 2021 (1). It is characterized by a fast epidemic growth relative to the B.1.617.2 (Delta) variant (2). Several epidemiological factors may contribute to the fast relative growth rate of this new variant. Firstly, immune evasion (3-5). Secondly, higher intrinsic transmission potential (6) (an increase in the basic reproduction number, defined as the average number of secondary cases generated by an infectious individual in a susceptible population). Thirdly, a shorter serial interval (i.e., the duration of time between symptom onset of a case and its infector). A variant with a shorter serial interval as compared to another variant with the same reproduction number, would have an increased epidemic growth rate. Whereas early reports provide evidence for substantial immune evasion and suggest an increased transmission potential (3-6), little is known about the serial interval of the Omicron variant. We assess whether the serial intervals of the Omicron BA.1 and Delta variant differ by comparing transmission pairs of both variants during the same time period.

The Omicron BA.1 variant was first identified in the Netherlands in a case whose sample was obtained on November 19, 2021. Symptom onset dates and postal codes of diagnosed SARS-CoV-2 cases are reported to a national surveillance database. If an infector of the case has been identified through source and contact tracing, a unique identifier of this infector is reported as well. We identified pairs of a primary case and a secondary case from this national surveillance database and measured the serial interval as difference between symptom onset day of a case and its infector.

A fraction of the cases reported in the national surveillance database were tested in two laboratories that analyze specimens with the TaqPath COVID-19 RT-PCR Kit (ThermoFisher Scientific). This PCR kit targets three genes. Failure of the probe targeting the S-gene, while the Orf1ab and N probe result in a proper signal (S-gene target failure (SGTF), also referred to as S-dropout), identifies the presence of a deletion in the S-gene (spike amino acid residues Δ69–70) which has been associated with the Omicron BA.1 variant but not with Delta. Non-SGTF is highly predictive of the Delta variant and SGTF is highly predictive for the Omicron BA.1 variant during times with little to no circulation of other variants (4). With lower viral loads, SGTF allocation is less accurate as the S-gene target is the least sensitive target of the three genes. Therefore, a stringent threshold of ≤ 30 cycle threshold (Ct) values were used on the Orf1ab and N targets for inclusion in further analyses.

We included transmission pairs with a minimum serial interval of -5 days and a maximum serial interval of 15 days. We included transmission pairs with a symptom onset date for the infector between 13 and 26 December 2021 (week 50 and 51), as reported by 24 January 2022, and report our results by week. The overall share of Omicron variant BA.1 detected in test positive cases was 9.0% in week 50 and 28.6% in week 51 (7). A cohort approach was followed to minimize the impact of data truncation and differences in epidemic growth by variant on the outcome. Analysis showed that most cases were tested and reported within five days after symptom onset. Combined with a maximum serial interval of 15 days, this would mean that a secondary case of the cohort would be tested and reported at the time the data were retrieved from the notification system. To ensure independent serial intervals, we included only unique infectors by choosing one of their cases at random. We excluded transmission pairs where infector or case had a missing postal code, where both infector and case lacked SGTF results, or where infector and case had differing SGTF results.

We will refer to transmission pairs with an SGTF case or an SGTF infector as SGTF transmission pairs, and to transmission pairs with a non-SGTF case or a non-SGTF infector as non-SGTF transmission pairs. We will refer to transmission pairs with a case and infector with the same postal code as within-household transmission pairs, and to transmissions with a case and infector with a different postal case as between-household transmission pairs, because 97% of transmission pairs with identical postal code live within the same household (8).

In week 50 (13-19 December 2021) we identified 235 SGTF transmission pairs and 919 non-SGTF transmission pairs, excluding 14 pairs with opposing SGTF results, 8 pairs without postal code, 193 pairs with non-unique infectors, and 6 pairs with a serial interval outside the range of -5 to 15 days. The mean serial interval of 3.5 days (sd 2.4 days) for the 164 SGTF within-household pairs was significantly shorter than the mean serial interval of 4.1 days (sd 2.8 days) for the 761 non-SGTF within-household pairs (Fig 1, bootstrapped p-value = 0.0026). A similar but not significant difference was found between the mean serial interval of 3.3 days (sd 2.4 days) for the 71 SGTF between-household pairs and the mean serial interval of 3.5 (sd 2.8 days) days for the 158 non-SGTF between-household pairs (bootstrapped p-value = 0.24). We grouped the within-household transmission pairs by the vaccination status of the infector and the case, and found that for each group the mean serial interval for SGTF transmission pairs is smaller than for non-SGTF transmission pairs, with statistical significance of this difference only when the infector is fully vaccinated (Fig S1).

**Figure 1.**
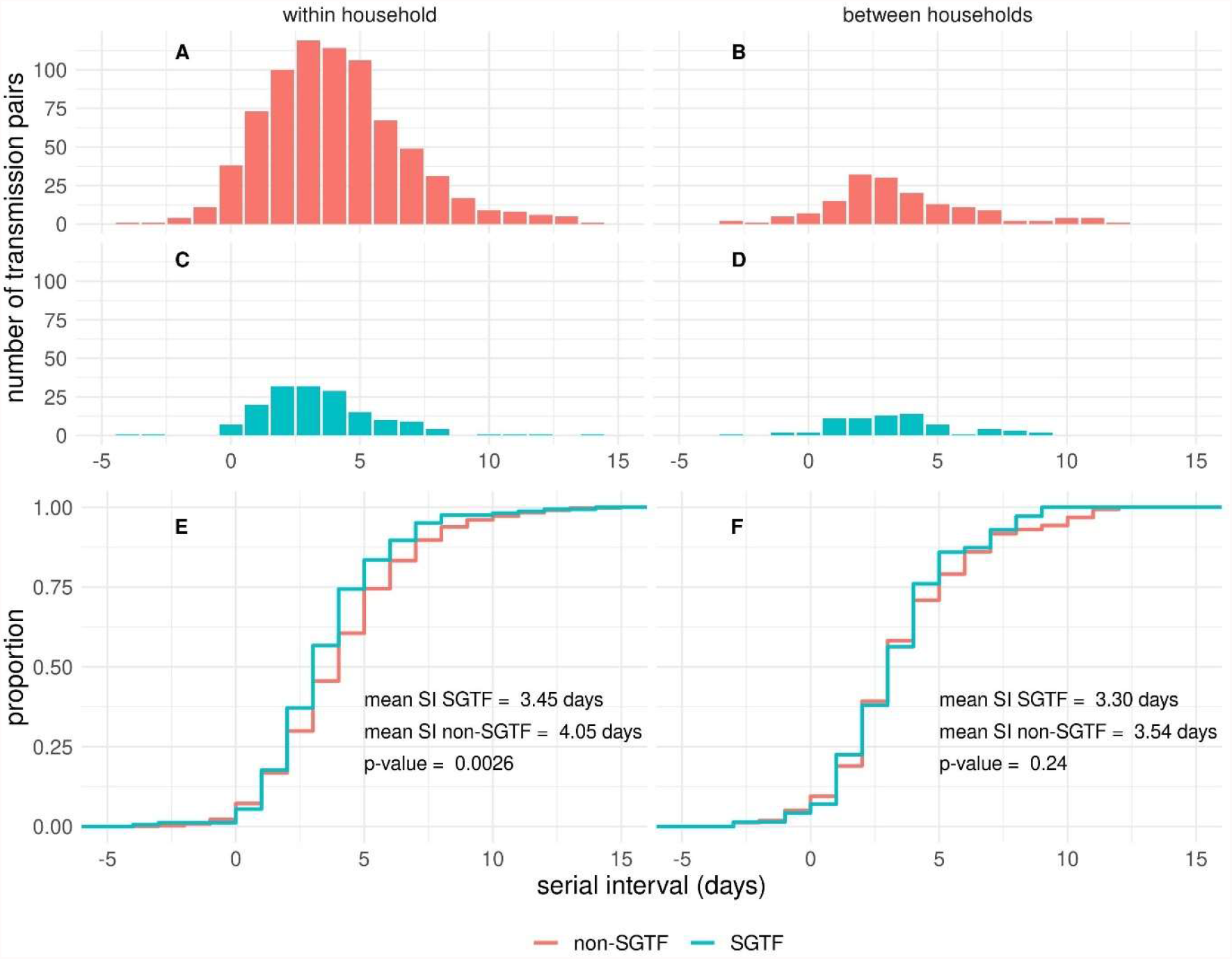
Observed distribution of serial interval of SARS-CoV-2 transmission pairs with infectors having their symptom onset date in week 50 (13 - 19 December 2021) in The Netherlands, (A) for non-SGTF within-household transmission pairs, (B) non-SGTF between-household transmission pairs, (C) SGTF within-household transmission pairs, (D) SGTF between-household transmission pairs, and the difference between the empirical cumulative density functions for SGTF and non-SGTF transmission pairs (E) within households and (F) between households (data in Tab S1). Significance of having different mean serial intervals is tested by bootstrapping.

In week 51 (20-26 December 2021) we identified 673 SGTF transmission pairs and 702 non-SGTF transmission pairs, excluding 13 pairs with opposing SGTF results, 9 pairs without postal code, 239 pairs with non-unique infectors, and 2 pairs with a serial interval outside the range of -5 to 15 days. Also in this week, the mean serial interval of 3.0 days (sd 2.3 days) for the 480 SGTF within-household pairs was shorter than the mean serial interval of 3.2 days (sd 2.6 days) for 572 non-SGTF transmission pairs (bootstrapped p-value = 0.084). A slightly shorter serial interval for SGTF versus non-SGTF was observed for between-household pairs (Fig S2).

In addition to the transmission pairs, we studied cases with known exposure information that allows to infer the incubation period (9). We identified 258 SGTF cases and 255 non-SGTF cases with reported symptom onset date between 1 December 2021 and 2 January 2022 (i.e., 13% of all cases with known exposure information in that period). The median incubation period is estimated to be 2.8 days for SGTF cases and 4.0 days for non-SGTF cases, with non-overlapping credible (Fig S3 and Tab S2).

This early investigation offers evidence to support a shorter serial interval among the SGTF transmission pairs presumed to be caused by the Omicron variant as compared to the non-SGTF transmission pairs presumed to be caused by the Delta variant. This lends support to the hypothesis that the recent rapid growth of the Omicron BA.1 variant is in part driven by a shortened serial interval as compared to infections with the Delta variant. The observed difference of 0.2 – 0.6 days is in line with the difference in the incubation period between the two variants.

During the study period the contact tracing guidelines differed for the two variants regarding contacts outside the household. Until 23 December 2021 guidelines for the Omicron variant were stricter than for Delta infections, with longer isolation and quarantine periods and requiring quarantine also for immune contacts. These differences might offer a possible explanation for the observed shorter serial interval of the SGTF transmission pairs between households compared to within households. However, these differences do not explain for the observed shorter serial interval of the SGTF transmission pairs within households and the shorter incubation period of SGTF cases. Therefore, the observed difference in between-household pairs is also expected to be due to the difference in variants.

To generalize the observed differences between serial interval for SGTF and non-SGTF transmission pairs, proper control for the control measures in place and other confounding factors such as age and vaccination status of the case and its infector are required. The difference in the duration of serial interval between successive weeks suggest a possible effect of changing control measures (stricter measures were implemented on 19 December 2021) and of subsequent changing behaviour, including increased testing behaviour around the Christmas period.

The reported values of the mean serial interval of 3.5 and 3.0 days for the Omicron variant are a bit longer compared to earlier reported tentative estimates. Mean serial intervals of 2.2 days and 2.8 days (range 1-7 days) were reported for an outbreak in South Korea (10, 11). Kremer et al. (manuscript to be published on medRxiv) report a mean serial interval of 2.75 days for the Omicron variant and 3.00 days for the Delta variant in Belgium. These values are more in line with the mean serial interval of 3.0 days that we observed for the period of 20 – 26 December 2021 in the Netherlands. The reported value of the median incubation period of 2.8 days for the Omicron variant is in line with earlier estimates. A median incubation period of 3 days for Omicron was reported for a superspreading event in Norway (12) and for a cluster in Nebraska (13). Although not all of these earlier reports allowed for a direct comparison between the reported values for the mean serial interval and the median incubation period between the Omicron and Delta variant in the same period, the reported low values suggest that also in these settings the serial interval and the incubation time of the Omicron BA.1 variant are shorter than those for the Delta variant.

There are indications for a different place of replication and a different route of entry for the Omicron variant, which suggests a mechanism to account for a shorter serial interval and a shorter incubation period (6, 14). Further studies that include the viral load and shedding dynamics relative to the symptom onset date of the primary case are crucial.

A short serial interval offers, next to immune evasion and higher transmissibility, an explanation for the growth advantage of the Omicron BA.1 variant over the Delta variant. A short serial interval and a short incubation period will make timely contact tracing more challenging, which will have a negative impact on reducing onward transmission (15). Mitigating the observed rapid spread of the new virus variant will therefore continue to require multi-layered interventions such as case finding and contact tracing, as well as booster vaccination and non-pharmaceutical interventions.

## Data Availability

All data produced in the present work are contained in the manuscript

## Supplementary material

### Data

**Table S1.**
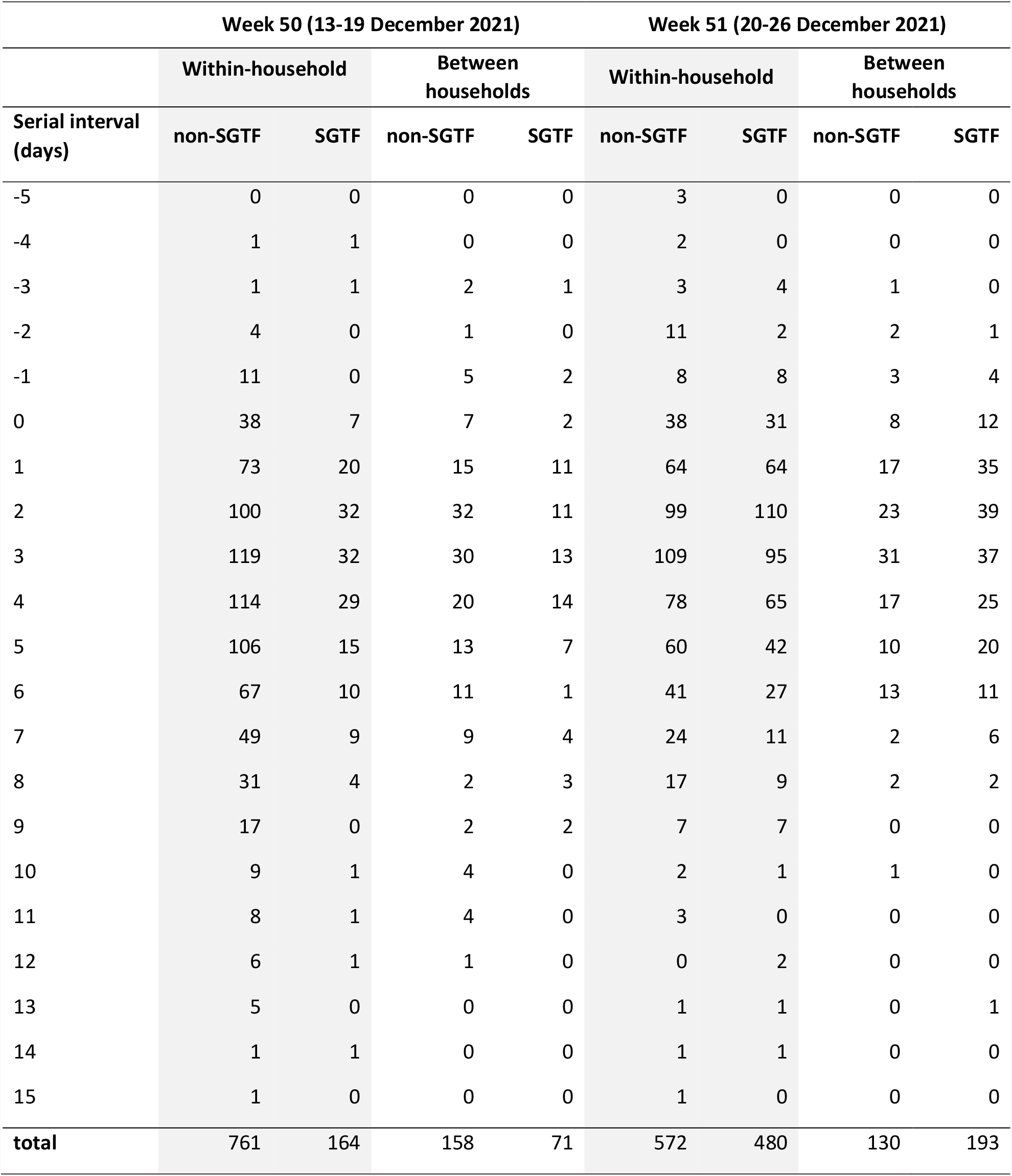
Number of observed transmission pairs by serial interval, study week, type (within or between households) and SGTF result; data shown in Figure 1 in main text for week 50 and in Figure S2 for week 51.

| Serial interval<br>(days) | Week 50 (13-19 December 2021) |  |  |  | Week 51 (20-26 December 2021) |  |  |  |
| --- | --- | --- | --- | --- | --- | --- | --- | --- |
|  | Within-household |  | Between households |  | Within-household |  | Between households |  |
|  | non-SGTF | SGTF | non-SGTF | SGTF | non-SGTF | SGTF | non-SGTF | SGTF |
| -5 | 0 | 0 | 0 | 0 | 3 | 0 | 0 | 0 |
| -4 | 1 | 1 | 0 | 0 | 2 | 0 | 0 | 0 |
| -3 | 1 | 1 | 2 | 1 | 3 | 4 | 1 | 0 |
| -2 | 4 | 0 | 1 | 0 | 11 | 2 | 2 | 1 |
| -1 | 11 | 0 | 5 | 2 | 8 | 8 | 3 | 4 |
| 0 | 38 | 7 | 7 | 2 | 38 | 31 | 8 | 12 |
| 1 | 73 | 20 | 15 | 11 | 64 | 64 | 17 | 35 |
| 2 | 100 | 32 | 32 | 11 | 99 | 110 | 23 | 39 |
| 3 | 119 | 32 | 30 | 13 | 109 | 95 | 31 | 37 |
| 4 | 114 | 29 | 20 | 14 | 78 | 65 | 17 | 25 |
| 5 | 106 | 15 | 13 | 7 | 60 | 42 | 10 | 20 |
| 6 | 67 | 10 | 11 | 1 | 41 | 27 | 13 | 11 |
| 7 | 49 | 9 | 9 | 4 | 24 | 11 | 2 | 6 |
| 8 | 31 | 4 | 2 | 3 | 17 | 9 | 2 | 2 |
| 9 | 17 | 0 | 2 | 2 | 7 | 7 | 0 | 0 |
| 10 | 9 | 1 | 4 | 0 | 2 | 1 | 1 | 0 |
| 11 | 8 | 1 | 4 | 0 | 3 | 0 | 0 | 0 |
| 12 | 6 | 1 | 1 | 0 | 0 | 2 | 0 | 0 |
| 13 | 5 | 0 | 0 | 0 | 1 | 1 | 0 | 1 |
| 14 | 1 | 1 | 0 | 0 | 1 | 1 | 0 | 0 |
| 15 | 1 | 0 | 0 | 0 | 1 | 0 | 0 | 0 |
| **total** | **761** | **164** | **158** | **71** | **572** | **480** | **130** | **193** |

### Serial interval of within-household transmission pairs in week 50 (13 - 19 December 2021), by vaccination status

**Figure S1.**
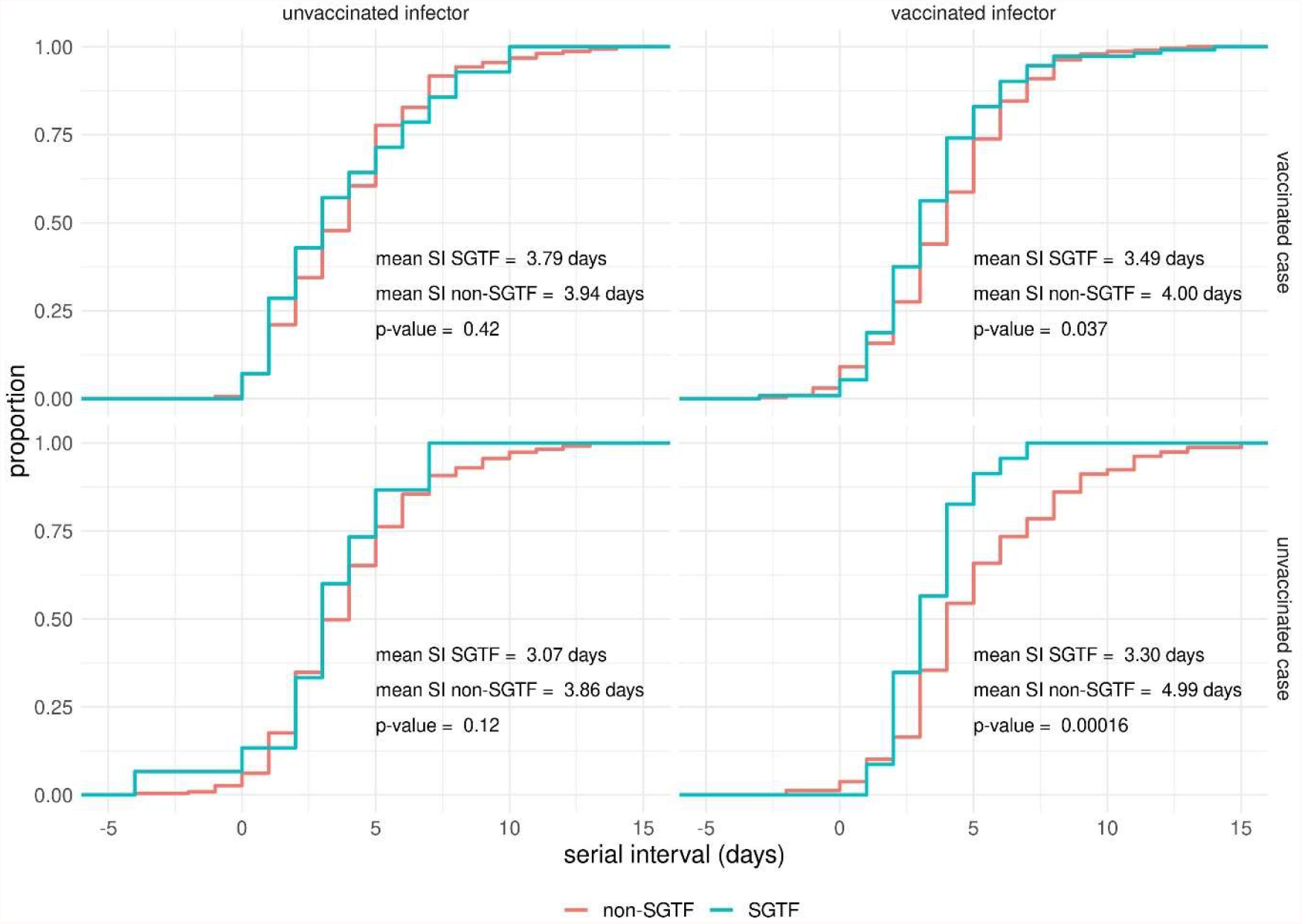
Observed distribution of serial interval of SARS-CoV-2 within-household transmission pairs with infectors having their symptom onset date in week 50 (13 - 19 December 2021) in The Netherlands, by vaccination status of the infector and case. A vaccinated status encompasses fully vaccinated persons including those who received a booster vaccination; an unvaccinated status encompasses unvaccinated and partially vaccinated persons. The panels show the difference between the empirical cumulative density functions for SGTF and non-SGTF transmission pairs by vaccination status of the index (columns) and case (rows). Significance of having different mean serial intervals is tested by bootstrapping.

### Serial interval for week 51 (20 – 26 December 2021)

**Figure S2.**
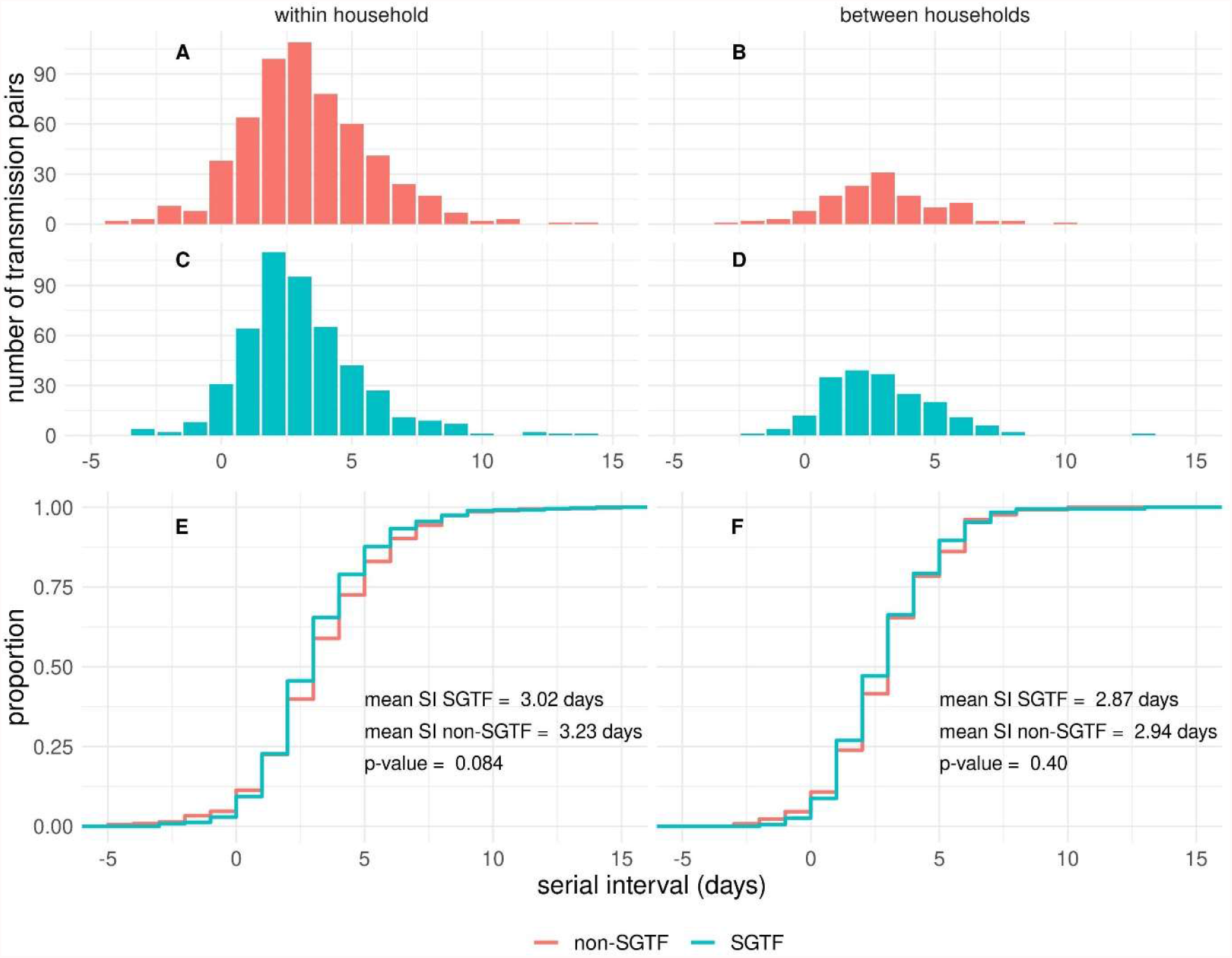
Observed distribution of serial interval of SARS-CoV-2 transmission pairs with infectors having their symptom onset date in week 51 (20 - 26 December 2021) in The Netherlands, (A) for non-SGTF within-household transmission pairs, (B) non-SGTF between-household transmission pairs, (C) SGTF within-household transmission pairs, (D) SGTF between-household transmission pairs, and the difference between the empirical cumulative density functions for SGTF and non-SGTF transmission pairs (E) within households and (F) between households (see Appendix for data). Significance of having different mean serial intervals is tested by bootstrapping.

### Incubation period

For 258 SGTF cases and 255 non-SGTF cases, with a symptom onset date between 1 December 2021 and 2 January 2022, the exposure window and symptom onset are reported. The parameters are estimated using a previously published method (Backer et al., Euro Surveill., 2020). A Weibull distribution was found to best fit the data compared to gamma and lognormal distributions. The incubation period for SGTF cases are shorter than for non-SGTF cases and the credible intervals are non-overlapping (Fig. S3 and Tab. S2).

**Figure S3.**
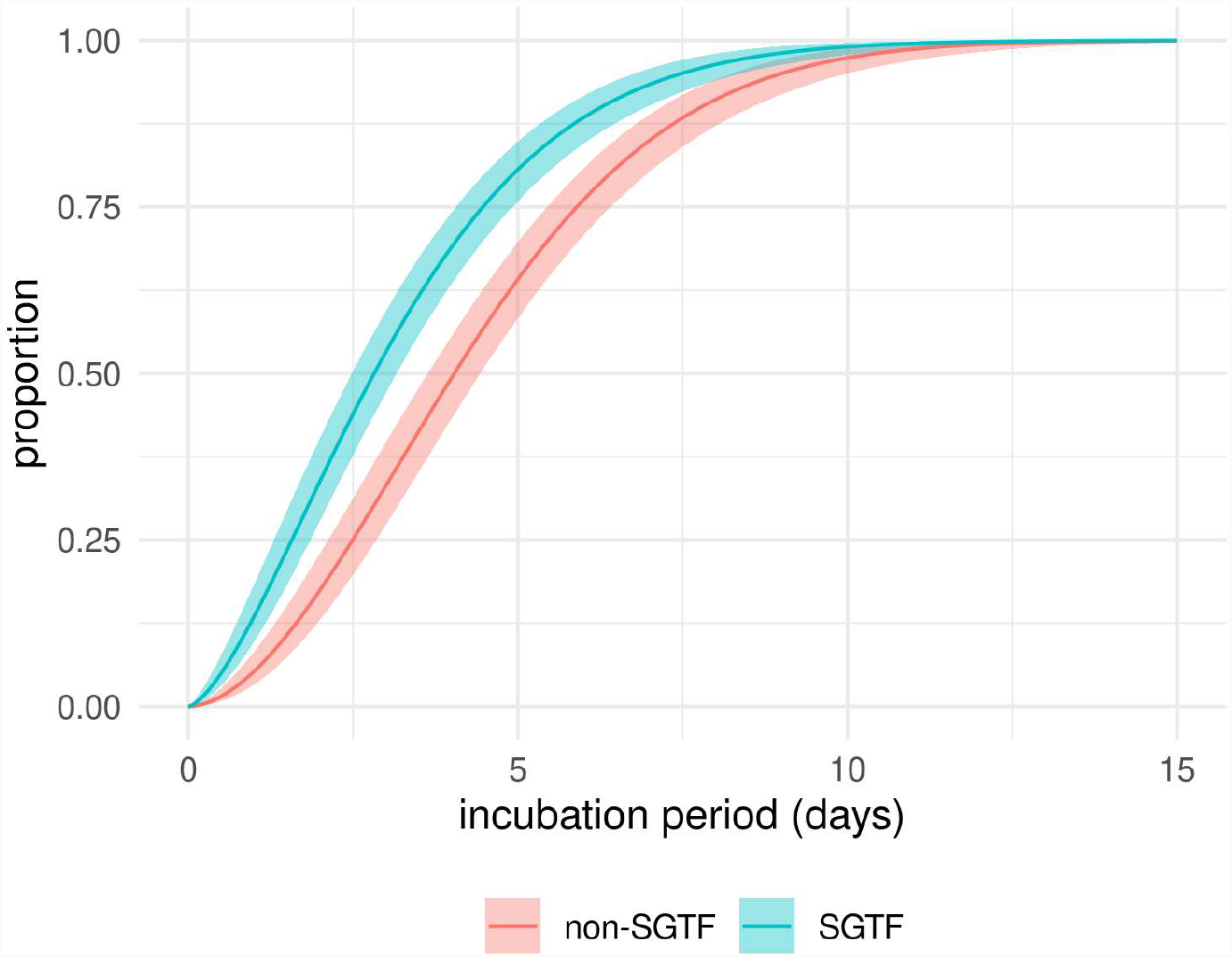
Estimated cumulative density function of the incubation period for 258 SGTF cases and 255 non-SGTF cases, with a symptom onset date between 1 December 2021 and 2 January 2022.

**Table S2.** Posterior parameter estimates for Weibull distributed incubation period

|  | non-SGTF |  | SGTF |  |
| --- | --- | --- | --- | --- |
|  | median | 95% cred. int. | median | 95% cred. int. |
| Scale parameter | 4.93 | 4.51 – 5.37 | 3.60 | 3.23 – 3.98 |
| Shape parameter | 1.83 | 1.59 – 2.08 | 1.50 | 1.32 – 1.70 |
| Mean (days) | 4.4 | 4.0 – 4.8 | 3.2 | 2.9 – 3.6 |
| Median (days) | 4.0 | 3.6 – 4.4 | 2.8 | 2.5 – 3.2 |

